# Personalizing mobile applications for health based on user profiles: A preference matrix from a scoping review

**DOI:** 10.1101/2025.04.22.25326205

**Authors:** Laëtitia Gosetto, Gilles Falquet, Fréderic Ehrler

## Abstract

The World Health Organization identifies unhealthy behaviors, such as smoking, as significant risk factors contributing to mortality and morbidity, underscoring the necessity to adopt healthier habits. The increasing prevalence of health applications (apps) presents opportunities for promoting healthier lifestyles. Notably, personalized mobile health (mHealth) interventions can enhance user engagement and their effectiveness. Our scoping review aims to contribute to guide the personalization of mHealth interventions for health behavior change by defining which mechanisms should be favored for a given user profile. Online databases were searched to identify articles published between 2008 and 2024 describing the topic of personalization, behavior change apps and mobile app mechanisms. Of 1806 articles identified, 18 articles were retained. We then categorized the mechanisms and user profiles described in the selected articles into existing taxonomies. Finally, the relationship between the user profiles and mechanisms were reported. The four user profiles identified included personality and gamer profiles. Twenty-one mechanisms extracted from the articles were categorized as behavioral change techniques, gamification or mobile app mechanisms, with limited numbers of preference relations between mechanisms and user profiles. The relation matrix was not complete and covered only 51% of possible relations: game mechanisms, 30%; behavioral change techniques, 16%; and app mechanisms, 5%. Two user profiles, the Big Five (18%) and Hexad scale (20%), covered 38% of relations, whereas the two remaining user profiles contributed to the remaining 13%. Social mechanisms, including competition, cooperation and social comparison, exhibit strong connections to user profiles and are pivotal in persuasive system design. Self-efficacy theory links mechanisms such as self-monitoring, social persuasion and rewards to behavior change. However, only 51% of potential relationships between profiles and mechanisms were identified. Adapting mHealth content based on user profiles requires reliable personality assessments and privacy-conscious data collection to enable personalized, profile-specific interventions for improved outcomes.

**Author Summary:** The promotion of healthy behavior, as well as addressing health risk factors that contribute to mortality, such as sedentary lifestyles, has led to a proliferation of mHealth apps. These apps have the potential to facilitate behavior change and offer a variety of features, including reminders, progress tracking and personalized interventions, which have been demonstrated to enhance user engagement and adherence. Personalization is of critical importance in the process of adapting interventions to align with the specific characteristics and needs of individual user profiles. The use of tailored messages and feedback has been demonstrated to be more effective than the use of generic ones, particularly in the context of promoting physical activity and weight loss. The incorporation of game design elements is also a prevalent feature in health apps, with evidence suggesting that it positively impacts on user engagement and motivation. However, there is a lack of comprehensive frameworks that provide guidance on their implementation in mHealth interventions. Here, we aim to optimize the effectiveness of interventions designed to facilitate health behavior change by defining game mechanisms, behavior change techniques and app mechanisms employed to personalize apps based on user profiles.

## Introduction

Certain behaviors increase the risk factors for mortality. According to the World Health Organization, the five leading global risk factors contributing to mortality are hypertension (13% of deaths), smoking (9%), high blood glucose (6%), sedentary lifestyle (6%), and being overweight (5%) [1]. It is therefore vital to encourage individuals to adopt healthy behaviors, such as smoking cessation or regular exercise. For this reason, the number of mobile health (mHealth) applications (apps) entering the market with the objective to help individuals to adopt healthier lifestyles is increasing rapidly. As of June 2022, there were more than 318,000 health apps available [2].

Smartphone apps present novel opportunities to promote health-related behaviors by offering immediate access to health information, reminders for medication adherence, and support for tracking progress [3]. A systematic review of the literature demonstrated the efficacy of mobile apps in addressing a range of health concerns, including chronic diseases, blood sugar regulation and smoking cessation [4]. Similarly, they have been shown to be effective in promoting healthy eating behaviors [5]. Notably, reviews on the impact of mHealth on obesity [6,7] highlighted its efficacy in weight management, body mass index reduction, waist circumference and blood pressure improvement [6], thereby fostering desired behavior changes [7]. Most of these apps employ behavioral change techniques (BCT) to encourage healthy habits, with prevalent strategies encompassing self-monitoring, goal setting, feedback, social support and reminders [4,6,8–15].

These techniques are defined in taxonomies such as CALO-RE [16]. For example, self-monitoring includes recording specific behaviors, such as the maintenance of a food diary or tracking daily weight.

Nevertheless, it is challenging to use a single technique on an entire population as each individual possesses a particular user profile, which encompasses a multitude of characteristics that define an individual. These include demographic data (e.g., age, gender), personality traits (e.g., Big Five, Myers-Briggs Type Indicator), cognitive profiles (e.g., need for cognition, sensitivity to persuasion) and attitudes (e.g., strong engagement in health behavior). Therefore, personalizing the app according to the user’s characteristics is certainly relevant. By contrast, the term “customization” pertains to the act of adapting the content by the user themself. Furthermore, several reviews have highlighted the advantages of providing tailored content based on the user profile’s specifics (personalized messages with user name[17], tailored goals [15] or feedback [6]) to enhance the efficacy of BCT [6,15,17].

A substantial body of research has been dedicated to the topic of personalization using tailored messages. As evidenced by a meta-analysis, behavior change interventions are significantly more effective when tailored to demographic variables [17]. Moreover, apps designed to promote physical activity and weight loss, have been demonstrated to be more effective when using tailored goals and messages than generic ones [15]. Similarly, tailored feedback has been demonstrated to be an effective approach in the context of obesity apps [6]. A study by McCaroll et al. revealed that participants who received tailored text messages exhibited higher average weight loss compared to those who received standard messages [5]. In addition, personalization can encompass multiple determinants simultaneously, including demographics, medical variables and behavioral change theories. In a study personalizing pamphlets for breast cancer based on age, ethnicity, family history of breast cancer, access to insurance, breast cancer risk and constructs from the Health Behavior Preference matrix, individuals who received a tailored intervention expressed a greater intention to undergo screening than those who received standardized messages [18]. Furthermore, Jakob et al. demonstrated that personalization could not only increase user engagement and adherence to mHealth solutions [19], but also enhance the effectiveness of mHealth interventions [19,20]. Indeed, personalization affects the ’little e’ of engagement in the digital behavior change intervention (DBCI) defined by Cole-Lewis et al. This ’little e’ refers to the interaction the user has with the DBCI features, as well as with the behavior change intervention components/active ingredients specifically designed to influence behavior determinants. When users reach an optimal level of interaction with the digital tool and the behavior change components are appropriately aligned, the probability of achieving the desired behavioral outcome is increased [21].

Another method for enhancing the efficacy of mHealth in promoting behavioral change is through the integration of game elements. The concept of gamification is a widely employed method for facilitating behavioral change. The term “gamification” is defined as “the use of game design elements in non-game contexts” [22]. A review by Klock et al. addressed the issue of personalization, but in the context of user experience and user interface design in tailored gamification [23]. Of note, Lister et al. reported that 52% of health apps reviewed incorporated at least one gamification element [9]. This inclusion had a positive effect on the intention to use health apps, particularly among young individuals without health issues [12]. In the context of gamification apps, it is advisable to personalize the app according to user profiles. These profiles have been defined and refer to users’ preferences regarding specific gameplay styles, such as narrative or puzzle-solving.

Previous reviews on behavior changes using mHealth have predominantly concentrated on the assessment of the efficacy of diverse mechanisms for initiating health behavior modifications, such as feedback, gamification mechanisms or BCT [4,6,8,9,11–13]. However, despite also reporting the significance of personalizing text messages to enhance their efficacy [6,17], no framework has been proposed to direct the personalization of mobile apps for health behavior change, apart from one review - but in the context of gamification alone [23]. In light of the aforementioned limitations, our study aims to contribute to the field by providing a framework in the form of a preference matrix for the personalization of mHealth interventions for health behavior by delineating the mechanisms that are most effective for a given user profile.

## Methods

This scoping review follows the Preferred Reporting Items for Systematic reviews and Meta-Analysis extension for Scoping Reviews (PRISMA-ScR) checklist [24].

### Eligibility criteria

Inclusion criteria were articles published in English between 1 January 2008 and 31 December 2024 (with 2008 marking the creation of the App Store) in a journal, book chapter or conference proceeding and addressing the relationship between user profiles and BCT or mechanisms applied through mHealth. The user profile had to be a validated personality model. The mHealth intervention should not be restricted to a specific population (e.g., people suffering from cancer) in order to ensure the generalization of our framework. Exclusion criteria were articles pertaining to the personalization of information, such as recommendations or the personalization of medical procedures.

### Information sources

On 19 August 2024, we conducted a search of the following databases as a scoping exercise to identify relevant publications: Science Direct, Psycnet APA, ACM, PubMed, Springer, JSTOR, IEE, and Web of Science.

### Search

The following search terms were used and combined using Boolean operators and articles published after 2008 were filtered for ACM, PubMed, Psycnet APA, IEEE, Web of Science and Springer:

*(“Personalization” OR “Tailoring” OR “Adaptative” OR “Customization” OR “Individualization” OR “Personalized” OR “Contextualization” OR “User-specific” OR “Adaptation” OR “User-centric” OR “Gamification”) AND (“Personality” OR “Gamers profile” OR “Big-five” OR “Hexad scale” OR “brainHex” OR “need for cognition” OR “user characteristics” OR “User attributes” OR “User data” OR “User identity”) AND (“mHealth” OR “mobile app” OR “mobile application” OR “eHealth” OR “Digital Health” OR “Mobile health” OR “health app”) AND (“Behavior change” OR “Persuasive technology” OR “Behavior modification” OR “Behavioral adjustment” OR “Habit change” OR “Behavior transformation” OR “Attitude change” OR “Behavioral adaptation”)*.

As the search functionality of JSTOR, Science Direct and Springer does not permit queries of such a length, we conducted a search of these databases using the following query, which was filtered and limited to articles published prior to 2008:

*(“Personalization” OR Gamification) AND (“Personality”) AND (mHealth OR “mobile app” OR “eHealth” OR digital) AND (“behavior change”)*.

### Selection of sources of evidence

Based on the inclusion and exclusion criteria, an initial selection based on a scan of the titles and abstracts was made independently by three reviewers. The full texts of the retained articles were then screened by two reviewers to ascertain their eligibility. Any discrepancies in the selection of studies were resolved through a process of consensus and discussion.

Concurrently, additional articles were incorporated through a ‘snowballing’ process. We included articles that cited or were cited by the initially selected papers and that were not identified through our search. Full text articles had to fulfill the established criteria.

### Data charting process

As a first step, we undertook a comprehensive listing of all user profiles exhibiting preference relations to mechanisms identified in the selected articles. Subsequently, the aforementioned mechanisms were categorized according to the BCT taxonomy [13] or the game elements proposed by Werbach and Hunter [25]. Finally, a matrix was constructed to represent these relationships in tabular form.

### Data items

The variables extracted included those pertaining to the user profile model used (four user profiles: Big Five, BrainHex, Hexad scale and gender), the mechanisms with preference relations (Table 2), the personalization type (e.g., on game elements, app mechanisms) and the media type (e.g., mobile app, video game, communication).

**Table 1.** Characteristics and findings of the articles included in this review.

| Article author (year)/country | Type* | Aim of the articles | User profile model | Personalization type | Media |
| --- | --- | --- | --- | --- | --- |
| Anagnostopoulou, E., et al. (2017) [26] / Greece | EA | Examines the relation between persuasion, personality and mobility types in personalized mobility apps. | Big Five | Game elements and apps mechanisms | Mobile app and video game |
| Alqahtani, F. et al. (2022) [27] / Canada | EA | Explores the relationships between personality and features of a persuasive app for promoting mental and emotional well-being. | Big Five | Game elements, BCT mechanisms, and apps mechanisms | Mobile app |
| Altmeyer, M. et al. (2019) [28] / Germany | EA | Investigates Hexad user types and behavior change intentions as factors to personalize gamified, persuasive fitness systems. | Hexad scale | Game elements | Mobile app |
| Codish, D. et Ravid, G. (2014) [29] / Israel | EA | Highlights the potential influence that personality has on the perceived playfulness from gamification and ultimately the expected benefit from it. | Big Five | Game elements | Video game |
| Halko, S. et al. (2010) [30] / USA | EA | Explores the relation between personality and persuasive technologies in the context of health-promoting mobile apps. | Big Five | Game elements | Mobile app |
| Hallifax, S., et al. (2019) [31]/ France | EA | Investigates the preference of game elements according to the user's profile. | BrainHex, Big Five, Hexad scale | Game elements | Video game |
| Jia, Y., et al. (2016) [32]/ USA | EA | Investigates the relations among individuals' personality traits and perceived preferences for various motivational affordance used in gamification. | Big Five | Game element | Video game |
| Johnson, D., et al. (2012) [33]/ Australia | EA | Explores relation between personality, video game preference and gaming experiences. | Big Five | Game elements | Video game |
| Klock, A.C.T. et al. (2020) [23]/ Finland | R | Presentation of a standardized terminology of the game elements used in tailored gamification and the most suitable game elements used in tailored gamification. | Hexad scale, Big Five, BrainHex, | Game elements | Video game |
| Ndulue, C. et al. (2022) [34]/ Canada | EA | Investigates whether the effectiveness of persuasive strategies varies across two distinct domains (healthy eating and smoking cessation) for people of distinct personality traits. | Big Five | Game element | Mobile app |
| Mora, A., et al. (2019) [35]/ Spain | EA | Investigates user types and preferences for different game design elements. | Hexad scale | Game elements | Video game |
| Orji, R. (2014) [36] / Canada | EA | Investigates differences in persuadability and the perceived persuasiveness of behavior change strategies between genders. | Gender | Game elements, BCT mechanisms, and apps mechanisms | Video game |
| Orji, R., et al. (2017) [37] / Canada | EA | Investigates how different personalities respond to persuasive strategies that are used in persuasive health games and gamified systems. | Big Five | Game element | Video game |
| Orji, R., et al. (2014)[38]/ Canada | EA | Investigates the efficacy of persuasive strategies for different gamer types in serious games for health. | BrainHex | Game element | Video game |
| Orji, R. et al. (2018) [39]/ Canada | EA | Investigates how different gamification user types responded to 10 persuasive strategies depicted in storyboards representing persuasive gameful health systems. | Hexad scale | Game element, app mechanism and BCT mechanism | Video game |
| Tondello G., et al. (2017) [40]/ Canada) | EA | Describes the characteristics of the users who are more likely to enjoy each group of design elements in terms of their gender, age, gamification user type, and personality traits. | Hexad scale, Big Five | BCT mechanism and app mechanism | Video game |
| Tondello, G., et al. (2016) [41]/ Canada | EA | Presentation of the Hexad scale, a gamification user types' model. Presentation of the association between the Hexad user types with the Big Five. | Hexad scale, Big Five | Game elements | Video game |
| Tondello, G., et Nacke, L. (2020)[42] / Canada | EA | Validation of user preferences and effects of personalized gamification on task performance. | Hexad scale, Big Five | Game elements | Video game |

\*EA, experimental article; R, review.

**Table 2.** List of mechanisms with their definition and frequency of occurrence in the corpus.

| Mechanism | Frequency of occurrence in the corpus | Definition |
| --- | --- | --- |
| Rewards | 13 | Virtual rewards offered to users for engaging in the target behavior [45]. |
| Competition | 12 | Users can compete to accomplish the desired behavior [45]. |
| Cooperation | 12 | Users collaborate to achieve a shared objective [45]. |
| Collection | 12 | Allows users to gather virtual objects. |
| Progression | 11 | Users can track their progression with steps through the system's purpose over time, visualized with mechanisms like stars or flags along a path [24]. |
| Customization | 9 | In contrast to personalization—which involves adjusting automatically the system to the user—customization refers to the user's ability to modify the content or functionalities of the mobile application according to their own preferences [45]. This approach enables users to actively tailor the system based on users' choices. |
| Social support | 8 | Enables communication between users, such as through chat or sharing activities with other users [24]. |
| Challenge | 7 | Presents various situations that require effort from the user to be completed [24] (e.g., accomplishing 3 hours of physical activity per week). |
| Social comparison | 7 | An individual's perceptions of the prevailing beliefs and behaviors within a social group. |
| Self-monitoring | 5 | Users can track their behaviors, providing information on both past and current activities [45] |
| Avatar | 4 | Allows users to share their data in the system without revealing their name [24]. |
| Punishment | 4 | Virtually penalizes the user for not performing the desired behavior or reaching their goal [39]. |
| Prompt and cues | 3 | Usually a message delivered to the user to prompt or recall a behavior at a specified time, with the app or user defining when the message should be sent [48]. |
| Demonstration of the behavior | 3 | Enables users to observe the cause-and-effect linkage of their behavior, such as seeing a simulation of their bodies after a diet [45]. |
| Quest | 2 | Users can enter or define the objectives targeted for the activity they will perform [48]. |

### Synthesis of results

Articles were subjected to a three-stage analytical process. The first stage entailed the enumeration of all identified mechanisms and user profiles within the selected articles. Subsequently, taxonomies were sought that encompassed the retrieved mechanisms and profiles. The third stage involved regrouping the mechanisms according to the taxonomies. Finally, the interrelationships between the identified user profiles and the mechanisms were documented.

## Results

### Selection of sources of evidence

The literature search yielded a total of 1806 articles; 92 articles were selected based on their title and abstract. Among these, 56 were rapidly excluded by all reviewers following agreement, and 36 remained in disagreement between two reviewers. Following additional discussions between reviewers, 34/36 articles were excluded, and two were included among the articles selected based on their titles and abstracts. Following assessment of the full text of the articles, two met the established eligibility criteria. In addition, 16 articles were further incorporated into the review through the snowballing process. From the 18 articles, a list of mechanisms used to personalize interventions was extracted (Fig. 1).

**Fig 1.**
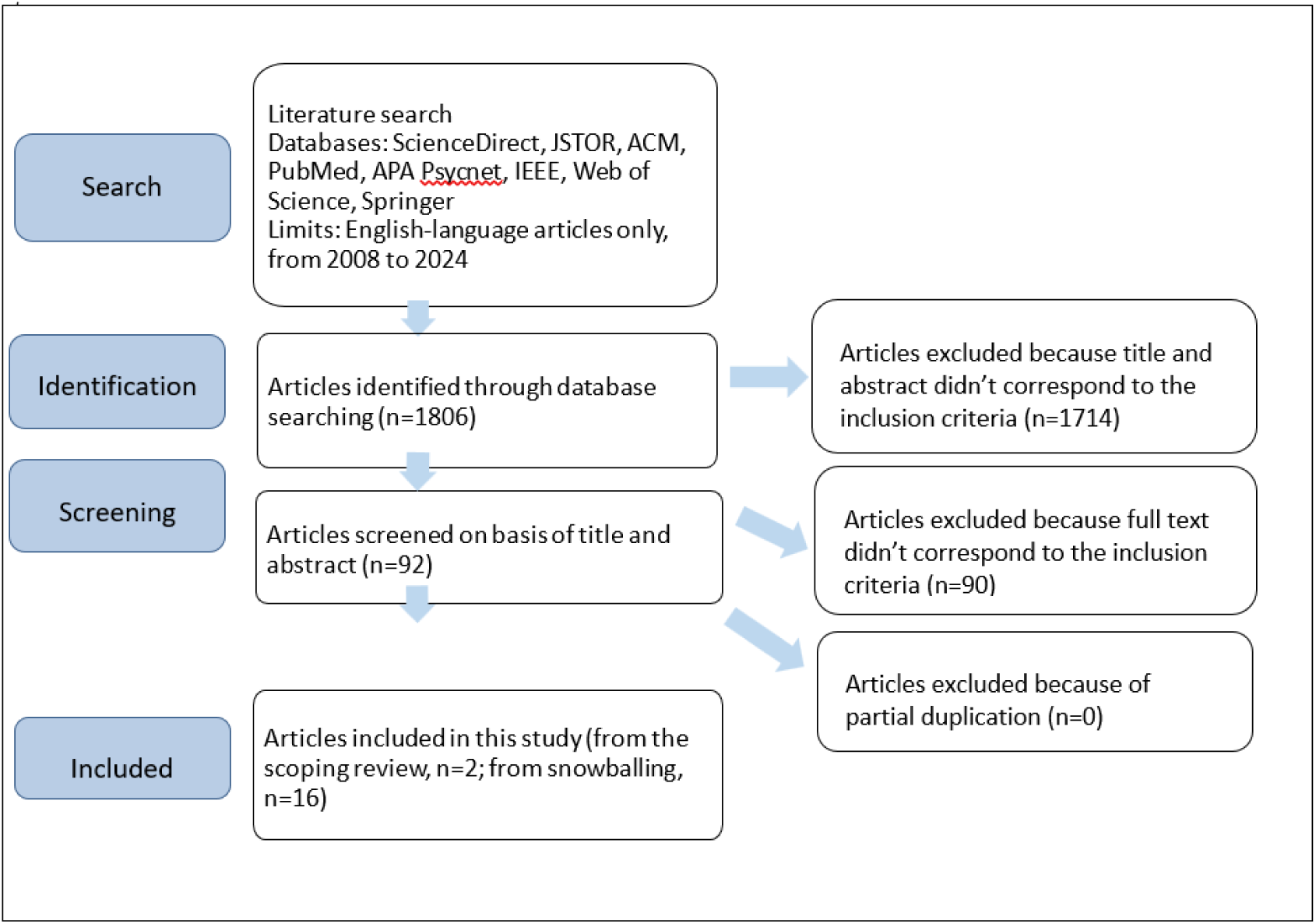
Source of evidence selection process.

### Characteristics of the sources of evidence

The characteristics of the 18 articles selected are presented in Table 1.

### Thematic presentation

#### User profiles

In the initial phase, we catalogued the user profiles identified in the literature. Five distinct types of user profile were identified, encompassing personality traits and player preferences. Personality was frequently assessed through various models, with the Big Five model being a prominent feature in 13/18 selected articles. These articles delineated personality based on five core dimensions: neuroticism, openness, conscientiousness, altruism and extroversion.

Derived from gamification theory, player profiles guide the process of mHealth personalization. The review revealed the existence of two distinct scales: BrainHex (featured in three articles) and Hexad scale (in 7 articles). Each scale defines player types and their preferred games or interactions, thereby offering insights into the gamification mechanisms that are favored by specific player archetypes. For example, according to the Hexad scale, philanthropic players who are motivated by goals and altruism respond well to collection and exchange elements within an app [44]. Finally, the demographic profile, which included gender data from a single article, was incorporated into the personalization profiles. Fig. 2 depicts the dimensions of the user profile.

**Fig 2.**
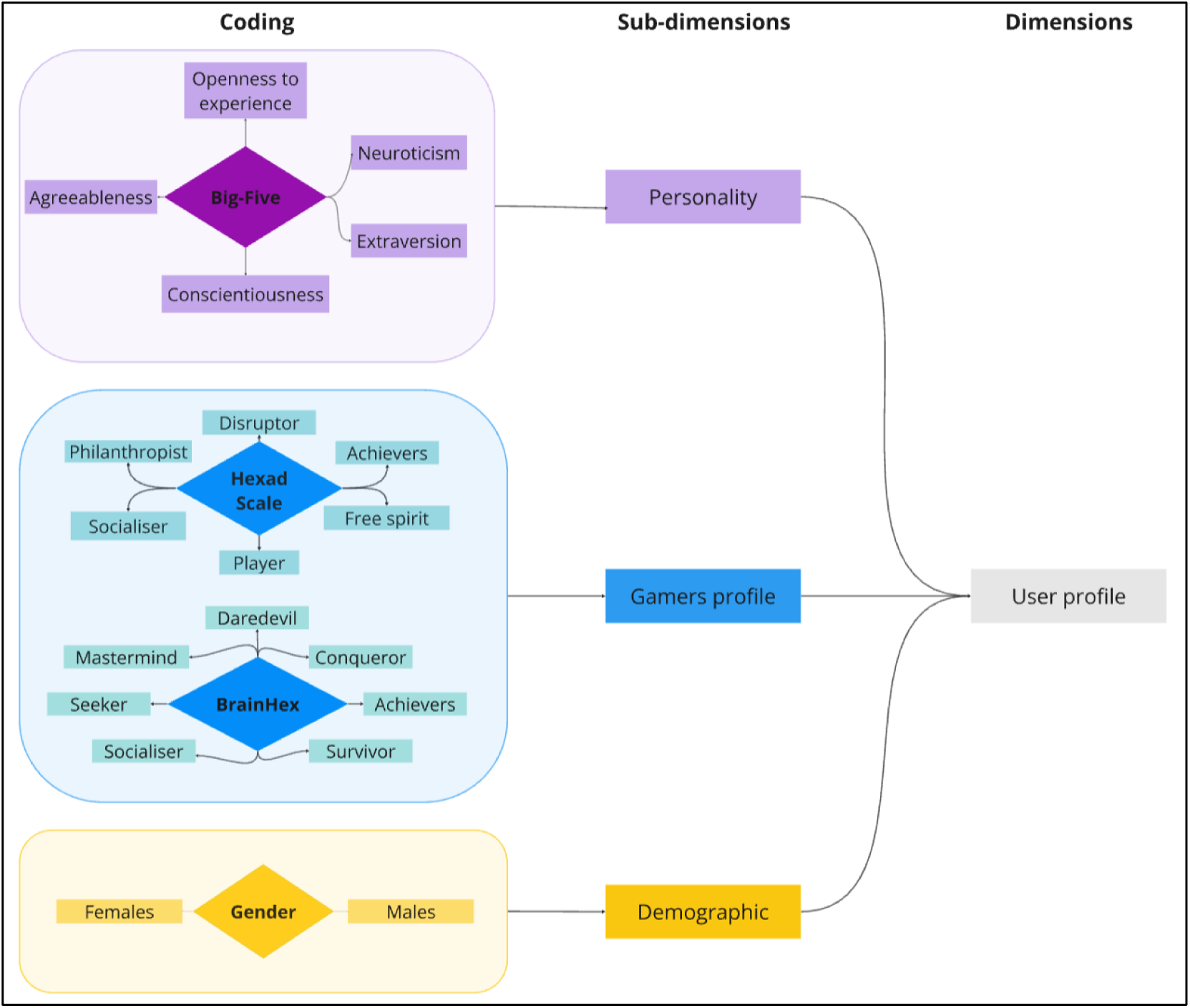
**Diagram showing the user profile dimensions.**

### Mechanisms

The literature revealed that 15 mechanisms have been studied in association with user profiles. In our context, the term ’mechanism’ is employed to denote the entirety of components that can be incorporated within an mHealth application with the objective of effecting behavioral change. This encompasses functionalities and gamification elements. Mechanisms are presented in order of their frequency of occurrence in the corpus (number of articles mentioning this mechanism), from the most to the least frequent (Table 2).

The aforementioned mechanisms have been classified into the following three sub-dimensions.

#### BCT taxonomy

A total of six mechanisms have been aligned with the Behaviour Change Taxonomy (BCT taxonomy). The authors define BCT as “observable, replicable and irreducible components of an intervention designed to alter or redirect causal processes that regulate behavior; i.e., a technique is proposed to be an ’active ingredient’ (e.g., feedback, self-monitoring, and reinforcement).” [16]. The BCT taxonomy is a standardized, hierarchically-structured classification of 93 distinct BCT, each with labels, definitions and examples. Its objective is to provide a reliable and consensus-based method for specifying, interpreting and implementing the active components of behavior change interventions across various disciplines and domains, including health and the environment. The following mechanisms have been classified within this taxonomy: social comparison, self-monitoring of behavior, demonstration of behavior, punishment, prompt and cues and social support.

#### Gamification classification

In order to categorize the specific mechanisms of games and gamification, we relied on the classification of game elements proposed by Werbach and Hunter [25]. In their analysis of over 100 gamification implementations, the researchers identified the most prevalent elements, which they termed the “Points, Badges and Leaderboards (PBL triad)”. The researchers then organized the remaining mechanisms into a category they termed ‘game elements’, defined as distinctive mechanisms inherent to video games. These game elements were subsequently classified into three principal categories pertinent to gamification: dynamics, mechanics and components, arranged in descending order of abstraction. *Dynamics* represent overarching aspects that are managed at a strategic level. *Mechanics* are fundamental processes that sustain engagement. *Components* are specific manifestations of mechanics or dynamics.

Eight mechanisms were aligned with the game element classification. *Progression* is classified as a dynamics category, while *competition, cooperation, challenge* and *rewards* are categorized as mechanics. The components category includes *avatars, collections*, and *quests*.

#### Mobile app mechanism

One mechanism did not align with both the game elements and the BCT taxonomy. The category of customization encompasses mechanisms that are specific to mobile apps.

For a graphical representation of the dimensions for the mechanisms, see Figure 3.

**Fig 3.**
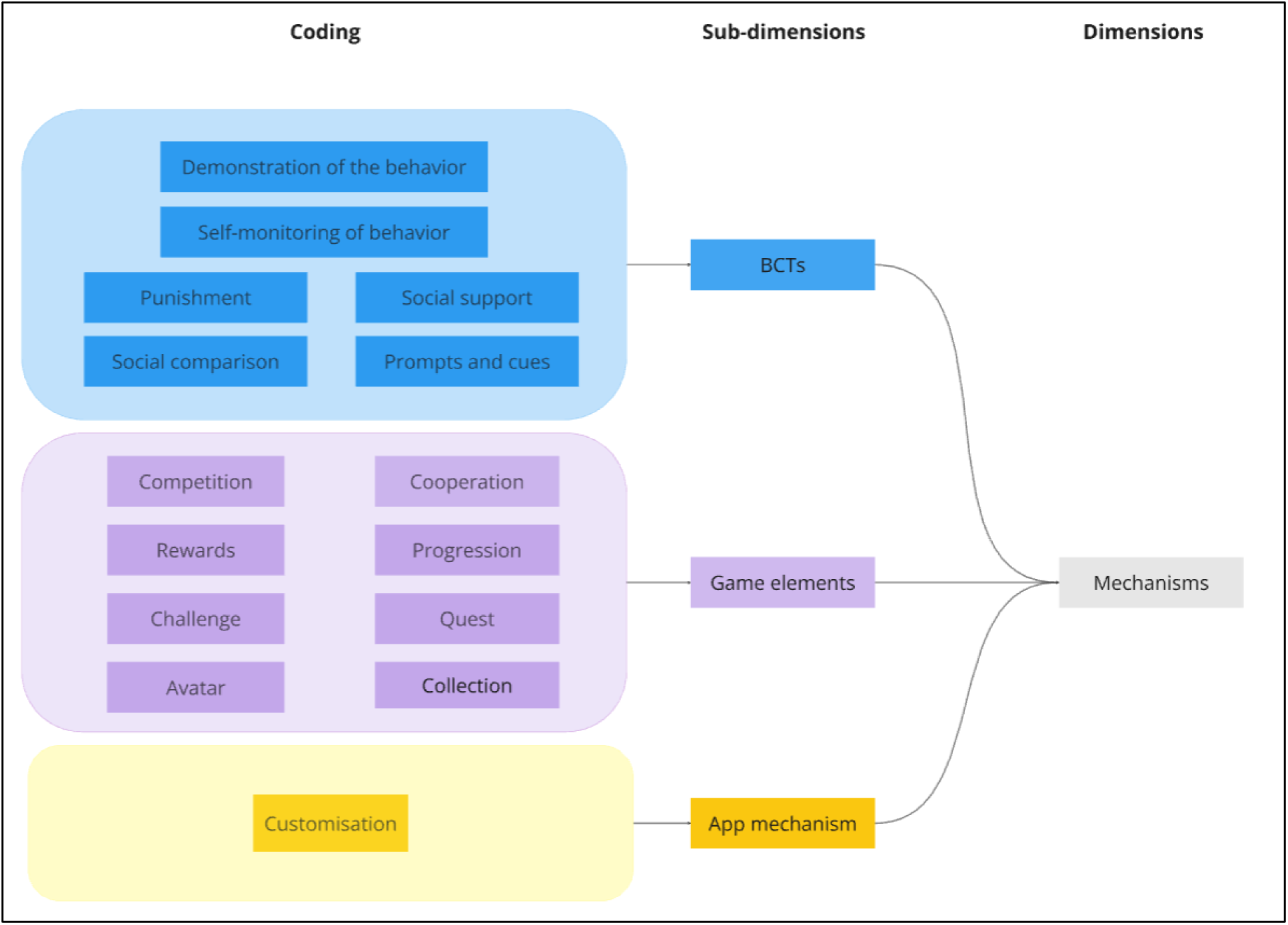
**Diagram illustrating the mechanisms’ dimensions.**

### Relations between dimensions

The selected articles identified and delineated the preference relations between the mechanisms and the user profiles. The aforementioned relations are summarized in Tables 3 and 4, with each relation referencing the corresponding article. Most preference relations were positive, indicating a preference for a specific mechanism within a given profile. Negative relations indicate a rejection of a mechanism (represented by an asterisk in Tables 3 and 4). It should be noted that the relation matrix is not yet complete and out of 300 possible relations, only 154 were identified in the corpus, representing 51% of the preference matrix.

**Table 3.** Relation between personality profiles and mechanisms.

|  |  |  | BCT mechanisms |  |  |  |  |  | Game elements |  |  |  |  |  |  |  | App mecha<br>nism |
| --- | --- | --- | --- | --- | --- | --- | --- | --- | --- | --- | --- | --- | --- | --- | --- | --- | --- |
|  |  |  | Prompts and cues | Demonstration of the behavior | Self-monitoring | Punishment | Social comparison | Social support | Progression | Competition | Cooperation | Collection | Rewards | Quest | Challenge | Avatar | Customization |
| Personality profile | Big Five | Openness to experience |  |  | [27, 37] | [37] | [37] | [27, 36] |  | [36, 37, 42]<br>[30]* | [31, 37]<br>[34]* | [36, 42] | [30, 37]<br>[34]* |  |  |  | [23, 37, 40] |
|  |  | Agreeableness | [27] | [37] | [26, 27, 37] | [36, 37] | [26, 37]<br>[31]* | [31, 38, 27] | [36, 38] | [34, 37]<br>[30, 31]* | [26, 34, 37] | [29, 42] | [26, 27, 34, 37] |  | [32, 38] |  | [26, 27, 37] |
|  |  | Conscientiousness |  | [37] | [37]<br>[26]* |  |  |  | [32] | [30, 29] | [30, 34]* |  | [29, 36] | [37] | [33] |  | [27]* |
|  |  | Extraversion | [27] | [37] | [27, 37] | [37] | [37, 23, 32] | [23, 27] | [23, 32, 36, 38] | [32, 37, 23] | [34, 37] | [23] | [27, 37, 23, 32, 30] | [37] | [33, 38] | [36] | [27, 37, 23]<br>[26]* |
|  |  | Neuroticism |  |  | [26] |  | [26] | [27] | [23] | [26]<br>[34]* | [30] | [23, 40] | [23, 26, 27, 34] |  |  |  | [26] |
| Demographic profile | gender | Females |  | [36] |  |  |  |  |  |  | [36] |  | [36] |  |  |  | [36] |
|  |  | Males |  |  |  |  |  |  |  |  |  |  |  |  |  |  |  |

\*For inverse relations.

**Table 4.**
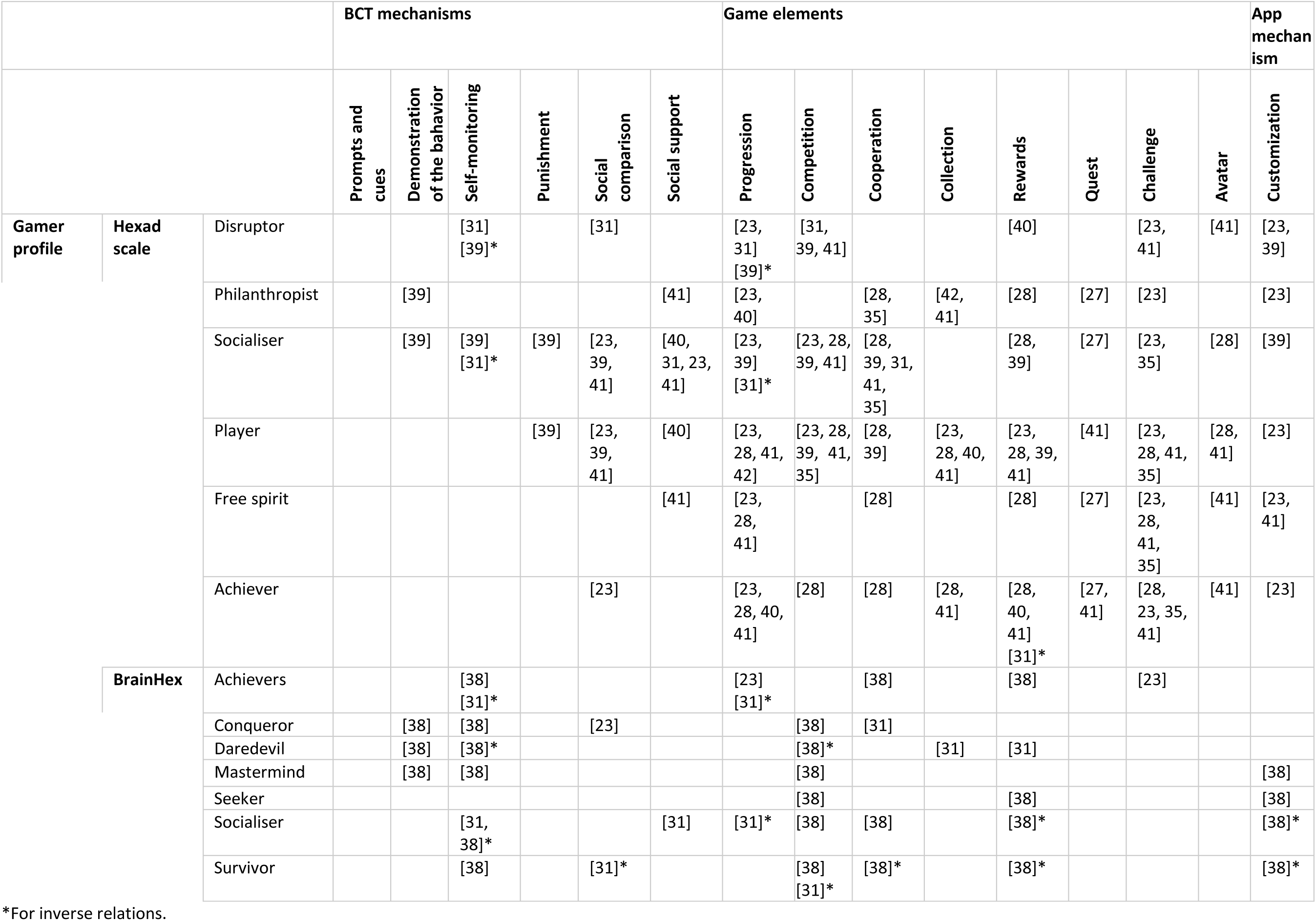
Relation between gamer profiles and mechanisms.

Our findings showed that two user profiles demonstrated a greater number of relations, i.e., the Big Five, representing 18% (55/300) of relations, and the Hexad scale comprising 20% (59/300) of relations (Table 3). In comparison, the BrainHex comprised 12% (35/300) of relations, while the gender comprised only 1% (4/300) of relations. The BCTs representing 16% (48/300) of relations, the Game elements 30% (90/300) relations, and app mechanism 5% (16/300) of relations.

Table 5 presents a classification of the number of relations per mechanism. The relative contributions of each mechanism to the preference matrix range from 1% to 6%. The mechanisms with the highest number of links are rewards, competition, customization and cooperation. It is evident that certain mechanisms exhibit relations with all traits present in a user profile. Rewards, competitions, customization, cooperation and self-monitoring demonstrate a connection with all five of the Big Five traits. Additionally, rewards, customization, progression and challenge exhibit a relationship with each of the Hexad scale traits. It has been demonstrated that other mechanisms exhibit strong relations with a user profile. Progression, social comparison, social support and collection have been found to be associated with four out of five Big Five traits. Furthermore, cooperation, quest, and avatar have been found to be associated with five out of six traits on the Hexad scale.

**Table 5.** Representation of the number of relations per mechanism on the preference matrix and the number of relations per mechanism per user profile.

| Mechanisms | Percentage total on the preference matrix (number of relations/total relation possible on the preference matrix) | Percentage on the Big Five (number of relations/total relation possible on the Big Five) | Percentage on gender (number of relations/ total relation possible on gender) | Percentage on the Hexad scale (number of relations/total relation possible on the Hexad scale) | Percentage on the BrainHex (number of relations/ total relation possible on the BrainHex) |
| --- | --- | --- | --- | --- | --- |
| Rewards | 6% (17/300) | 100% (5/5) | 50% (1/2) | 100% (6/6) | 71% (5/7) |
| Competition | 5% (16/300) | 100% (5/5) | 0% (0/2) | 67% (4/6) | 86% (6/7) |
| Customization | 5% (16/300) | 100% (5/5) | 50% (1/2) | 100% (6/6) | 57% (4/7) |
| Cooperation | 5% (15/300) | 100% (5/5) | 50% (1/2) | 83% (5/6) | 57% (4/7) |
| Self-monitoring | 4% (13/300) | 100% (5/5) | 0% (0/2) | 33% (2/6) | 86% (6/7) |
| Progression | 4% (12/300) | 80% (4/5) | 0% (0/2) | 100% (6/6) | 29% (2/7) |
| Challenge | 3% (10/300) | 60% (3/5) | 0% (0/2) | 100% (6/6) | 14% (1/7) |
| Demonstration of the behavior | 3% (9/300) | 60% (3/5) | 50% (1/2) | 33% (2/6) | 43% (3/7) |
| Social comparison | 3% (9/300) | 80% (4/5) | 0% (0/2) | 67% (4/6) | 29% (2/7) |
| Social support | 3% (9/300) | 80% (4/5) | 0% (0/2) | 67% (4/6) | 14% (1/7) |
| Collection | 3% (8/300) | 80% (4/5) | 0% (0/2) | 50% (3/6) | 14% (1/7) |
| Quest | 2% (7/300) | 40% (2/5) | 0% (0/2) | 83% (5/6) | 0% (0/7) |
| Avatar | 2% (7/300) | 20% (2/5) | 0% (0/2) | 83% (5/6) | 0% (0/7) |
| Punishment | 2% (5/300) | 0% (0/5) | 0% (0/2) | 33% (2/6) | 0% (0/7) |
| Prompts and cues | 1% (2/300) | 40% (2/5) | 0% (0/2) | 0% (0/6) | 0% (0/7) |

It is also worth noting that specific traits exhibit preference relations with nearly all mechanisms. Specifically: extraversion from the Big Five model exhibits relationships with all mechanisms; agreeableness from the Hexad model demonstrates relationships with 13/15 mechanisms; socializer from the Hexad model exhibits relationships with 13/15 mechanisms; and player from the Hexad model demonstrates relationships with 12/15 mechanisms. We can also observe that there is little relationship with gender. Only one article showed relations with gender, and only with women.

Finally, it was also observed that certain traits exhibit both preference and non-preference relationships for the same mechanism. For instance, the openness to experience trait exhibits a non-preference relationship with the cooperation mechanism, while the disruptor trait displays a non-preference relationship with the progression mechanism. This analysis reveals a total of 14 inconsistent relationships within the preference matrix.

## Discussion

In this study, a comprehensive review identified the preferred mechanisms for specific user profiles that can be employed in mobile apps to prompt behavioral change in health. The exploration revealed a variety of mechanisms including gamification mechanisms, mobile app mechanisms and BCT mechanisms. As a result, we have established connections between these user profiles and these mechanisms. A summary of the preferred and non-preferred mechanisms by user profile is provided in Table 6.

**Table 6.**
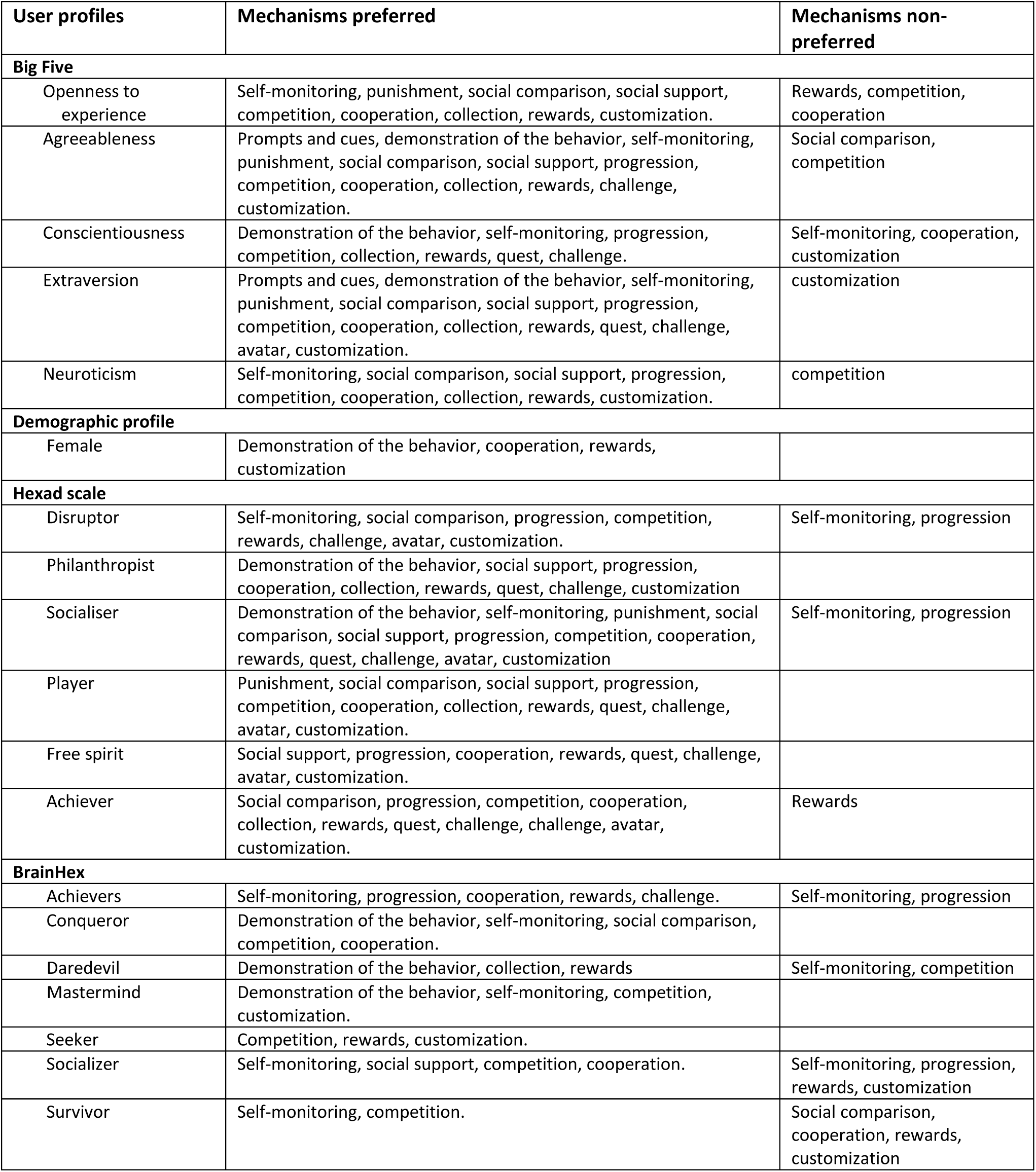
Summary of preferred and non-preferred mechanisms by user profile.

| User profiles | Mechanisms preferred | Mechanisms non-preferred |
| --- | --- | --- |
| **Big Five** |  |  |
| Openness to experience | Self-monitoring, punishment, social comparison, social support, competition, cooperation, collection, rewards, customization. | Rewards, competition, cooperation |
| Agreeableness | Prompts and cues, demonstration of the behavior, self-monitoring, punishment, social comparison, social support, progression, competition, cooperation, collection, rewards, challenge, customization. | Social comparison, competition |
| Conscientiousness | Demonstration of the behavior, self-monitoring, progression, competition, collection, rewards, quest, challenge. | Self-monitoring, cooperation, customization |
| Extraversion | Prompts and cues, demonstration of the behavior, self-monitoring, punishment, social comparison, social support, progression, competition, cooperation, collection, rewards, quest, challenge, avatar, customization. | customization |
| Neuroticism | Self-monitoring, social comparison, social support, progression, competition, cooperation, collection, rewards, customization. | competition |
| **Demographic profile** |  |  |
| Female | Demonstration of the behavior, cooperation, rewards, customization |  |
| **Hexad scale** |  |  |
| Disruptor | Self-monitoring, social comparison, progression, competition, rewards, challenge, avatar, customization. | Self-monitoring, progression |
| Philanthropist | Demonstration of the behavior, social support, progression, cooperation, collection, rewards, quest, challenge, customization |  |
| Socialiser | Demonstration of the behavior, self-monitoring, punishment, social comparison, social support, progression, competition, cooperation, rewards, quest, challenge, avatar, customization | Self-monitoring, progression |
| Player | Punishment, social comparison, social support, progression, competition, cooperation, collection, rewards, quest, challenge, avatar, customization. |  |
| Free spirit | Social support, progression, cooperation, rewards, quest, challenge, avatar, customization. |  |
| Achiever | Social comparison, progression, competition, cooperation, collection, rewards, quest, challenge, challenge, avatar, customization. | Rewards |
| **BrainHex** |  |  |
| Achievers | Self-monitoring, progression, cooperation, rewards, challenge. | Self-monitoring, progression |
| Conqueror | Demonstration of the behavior, self-monitoring, social comparison, competition, cooperation. |  |
| Daredevil | Demonstration of the behavior, collection, rewards | Self-monitoring, competition |
| Mastermind | Demonstration of the behavior, self-monitoring, competition, customization. |  |
| Seeker | Competition, rewards, customization. |  |
| Socializer | Self-monitoring, social support, competition, cooperation. | Self-monitoring, progression, rewards, customization |
| Survivor | Self-monitoring, competition. | Social comparison, cooperation, rewards, customization |

Personalized mobile health apps have been demonstrated to be more effective in inducing behavior change, particularly when the messages and goals are tailored to the individual rather than generic ones, as highlighted by Sporrel et al. [15]. Accordingly, the development of mobile apps should be aligned with the user profile. The literature reveals three distinct user profiles: personality (utilizing the Big Five model), player profile (comprising the Hexad scale and BrainHex) and gender. Each profile categorizes users based on a variety of criteria. For instance, the Big Five model can be used to distinguish a user who exhibits high levels of extraversion and low levels of altruism. It is also notable that a single profile may exhibit a preference or rejection for a given mechanism. For example, the disruptor profile may prefer progression [31] or may reject this mechanism according to another article [39]. This inconsistency in reports constitutes a minor proportion of the preference matrix (5%), yet it underscores the imperative for further investigation to resolve this discrepancy.

One review of the preference matrix addressed the concept of personalization within the context of gamification [23]. Of note, 71% of the relations presented in this review were validated by other articles in the corpus, especially with all the relations with a BrainHex profile. The BrainHex profile, a construct of particular interest, is comparatively underdocumented, with only three articles in the corpus directly addressing its relationship. This paucity of documentation may partially account for the absence of corroboration from our corpus. Further research is therefore necessary to gather more data related to BrainHex. Conversely, this review encompasses a mere 13% of the potential relationships with our matrix (39/300). This finding underscores the significance of our scoping review, which offers a more extensive array of content and establishes connections between user profiles and mechanisms.

### Social contact

A multitude of connections exist between the mechanisms associated with social contact and user profiles. It is noteworthy that social comparison, cooperation and competition, with 9, 15 and 16 relations, respectively, exceed the average number of relations per mechanism. This concept is of considerable significance, representing a category within the Persuasive System Design (PSD) framework that delineates the essential content and mechanisms for a persuasive system. Inclusion in the social support category of this framework further incorporates competition, cooperation and social comparison [43]. Notwithstanding meta-analyses that have affirmed the efficacy of interventions with a social network for behavioral change [33,44], social mechanisms are not frequently utilized in behavior change apps. Indeed, few apps in general include these social contact mechanisms. For example, of the 208 apps mechanism from Villalobos-Zuniga and Cherubini’s taxonomy, less than 20% are related to social contact [45]. However, it can be reasonably deduced that encouraging the integration of these mechanisms within the domain of mobile health technology would prove to be a valuable undertaking.

### Self-efficacy

A number of the most frequently retrieved mechanisms (with more than seven relations, representing the average number of relations per mechanism) can be associated with Bandura’s theory of self-efficacy [46], which has been demonstrated to influence both short- and long-term behavioral change [47]. The construct of self-efficacy is influenced by four primary sources: enactive attainment, vicarious experience, social persuasion and physiological/emotional states.

Enactive attainment entails observing one’s past performance and providing an accurate assessment of one’s abilities, which serves to encourage continued efforts. This is analogous to the self-monitoring mechanism, which enables individuals to evaluate their performance, such as by monitoring the number of daily steps. Furthermore, the self-monitoring mechanism is present in the PSD [43] under the dialogue support category and also in another scale, the App Behavior Change Scale [48].

Vicarious experience can be defined as the process of gaining confidence through observation of others who are perceived to possess similar abilities engaging in the behavior in question. This experience can be facilitated by various mechanisms, including social comparison, which enables users to compare themselves with others, and the social network, which allows the sharing of performance and progress with other users.

Social persuasion entails the utilization of verbal reinforcement to motivate individuals to act in a manner that is perceived to enhance their ability to succeed. The provision of social support enables users to receive verbal encouragement and engage in supportive conversations.

Physiological/emotional states are used to describe their impact at the time of success or failure on one’s sense of efficacy. A positive mood during a successful outcome leads to a more positive evaluation, whereas a negative mood during an unsuccessful outcome results in a lower sense of personal efficacy. It is therefore beneficial to cultivate a positive emotional state in users when they experience success. This can be achieved through the provision of rewards, such as badges or fictive coins.

### Relation between BCT taxonomy and game elements

In the process of categorizing mechanisms within the BCT or game elements, it became evident that certain mechanisms could be placed in both taxonomies. For example, competition may be classified as social comparison within the BCT, while rewards may also be subsumed into the BCT reward and threat category. In instances where a mechanism was identified in both taxonomies, we retained it exclusively within the game elements’ taxonomy. This decision was made in consideration of the comparison between gamification mechanisms and established behavioral change mechanisms that have been proven effective in digital health interventions [49].

### Persuasion strategies

The research conducted for this article revealed the potential for adapting Cialdini’s six principles of persuasion (*liking, reciprocity, consensus, commitment and consistency*, *authority, scarcity*) according to the personality of the user. These principles have been extensively applied in the domains of marketing and persuasive technology [50] and identify six methods for requesting compliance with a particular course of action. For example, the principle of liking posits that requests made by individuals with whom we have a positive affinity are more likely to be complied with [51], or consensus posits that individuals are inclined to replicate the actions of others who share similar characteristics with them. However, we have elected to exclude these principles from our framework on the grounds that they are not mechanisms in themselves, but rather a means of personalizing messages. It would be beneficial for future research to consider the personalization of messages according to user profiles in the context of mHealth. In particular, research has demonstrated that personalized messages based on the recipient’s personality are more effective [52].

### Need for cognition

Our research has been expanded to encompass additional user profiles, including those pertaining to Need for Cognition (NFC). Indeed, this concept has been the subject of ongoing interest for researchers in the field of psychology [50] with over 8000 citations in articles following the original NFC article of Cacioppo et Petty published in 1982 [53]. NFC is a construct that characterizes individuals based on their intrinsic motivation for engaging in cognitively demanding tasks [50].

However, the articles on this topic did not present any relations with the identified mechanisms, thus precluding their inclusion in the scoping review. Individuals with high NFC demonstrate a preference for messages with robust arguments, whereas those with low NFC exhibit no preference between strong and weak arguments [54]. Furthermore, the Elaboration Likelihood Model [55] posits that NFC may be linked to two distinct routes in the persuasion process. The central route pertains to the manner in which individuals pay attention to presented arguments, whereas the peripheral route involves the reliance on simple persuasive cues, such as the message’s source, when motivation or processing abilities are low. Consequently, individuals with low NFC may be inclined to utilize the peripheral route, whereas those with high NFC may demonstrate a proclivity for the central route [56]. It seems reasonable to suggest that the transmission of health behavior information and the formulation of feedback should be adapted on the basis of the user’s NFC level. However, no articles were identified that examined relationships according to this profile.

### Small number of relations

The articles selected for this review did not present a comprehensive account of the preference relations between all profile types and mechanisms. Indeed, 51% of potential relations were identified. This highlights an existing gap in knowledge although the limited number of articles included in this scoping review does not guarantee the comprehensive coverage of potential relations. Moreover, a considerable proportion of the studies included in this scoping review were conference proceedings rather than peer-reviewed journal articles. This prevalence may reflect the emerging nature of the field, but it also underscores a lack of robust, high-quality empirical research. The reliance on preliminary findings and non-archival sources highlights the urgent need for more rigorous, peer-reviewed investigations to establish a stronger evidence base in this area. It is also noteworthy that mHealth interventions identified in the literature rarely adapt their content based on user profiles. To identify additional relations, our search was expanded to include websites, video games, and various message types as these elements are commonly incorporated in mHealth.

### Applying the approach

The use of the preference matrix in practice would require the completion of two distinct steps. The initial step will be to define the user profile for the application. The second step will be to adapt the content to align with the identified profile.

The first step, identification of the user profile, presents a significant challenge. This process typically involves having users complete standardized tests (e.g., the Big Five Inventory 10 Item Scale [BFI-10], Revised NEO Personality Instrument [NEO-PI-R]) and subsequently making adjustments to the app based on the user’s scores. Nevertheless, this process has the potential to be time-consuming for the user. Some users may be reluctant to use the app if they are required to complete this type of questionnaire. Consequently, it is more rational to utilize automatic personality assessments. Research has demonstrated the feasibility of employing these assessments with existing data sources, including smartphone data (e.g., call duration, SMS, Bluetooth connection) [57,58], demographic data [59], social media [59–61], user activities on YouTube [62], language-based assessments [63], and wearable activity trackers [64,65]. However, such assessments necessitate the collection of data, which users often demonstrate reluctance to share, particularly when it comes to audio and video data [66]. Privacy concerns related to excessive data collection may results in users to perceiving behavioral changes influenced by these assessments in a negative light, which could ultimately diminish the perceived accuracy of the assessments [66].

Therefore, the most pragmatic approach is to make app personalization optional, allowing users to decide whether or not to engage with this functionality. Should the user express a desire for this functionality, they will be prompted to complete a brief personality questionnaire, utilizing the most concise version that has been empirically validated.

The second step would be to guarantee that the app contains solely the mechanisms that correspond to the dominant traits (which can be multiple) of the user’s profile in accordance with our preference matrix. For instance, for participants exhibiting a high score on the Big Five conscientiousness scale, the app would include demonstration of the behavior, self-monitoring and, if the app is gamified, the progression, competition, cooperation, collection, rewards, quest and challenge mechanisms. It is also possible to personalize according to several user profiles. These profiles may be characterized by the Big Five personality model, gender, or the Hexad scale.

Furthermore, it is possible to select mechanisms corresponding to a user’s dominant traits for each of these profiles.

This process can be initiated at the time of account creation within the app. As an alternative, the option may be presented subsequent to the creation of the account. Should the user wish to trial an application that has been tailored to their profile, they may do so.

### Limitations

The decision was taken to categorize the mechanisms in question in accordance with the BCT taxonomy [13] and the game elements defined by [25]. Other taxonomies, such as PSD [43], include also mechanisms such as rewards, social support and self-monitoring, but the BCT taxonomy was selected due to its extensive usage, with over 6000 citations. An alternative approach would have been to categorize the mechanisms according to the mechanisms of action (MoA), which appear to be a more relevant classification system [67]. However, the BCT taxonomy is more detailed and allows for a more precise level of granularity than the MoA, thereby enabling a more accurate classification of the mechanisms identified in the literature.

The classification of mechanisms into the taxonomy was conducted by a single researcher. To mitigate research biases and enhance the credibility and validity of the taxonomy, it is recommended that validation by other researchers, specifically investigator triangulation, should be employed. The process of triangulation serves to confirm the accuracy of the taxonomy classification, identify instances of conflicting classification, rectify errors and enhance the overall accuracy of the taxonomy [68]. In order to corroborate this classification, it would be beneficial to engage in dialogue with experts in the field, such as focus groups comprising various specialists (e.g., app developers, healthcare professionals, health psychologists, BCT experts, etc.).

Another limitation is the addition of a snowballing process, which makes the process less reproducible. Surprisingly, after performing the scoping review, some articles frequently cited in article introductions were not in our corpus. We therefore decided to add this snowballing process as these articles met our criteria, although they did not stand out among the results of our searches in the various selected journals.

Finaly, the reliability of the relations is questionable, as studies are conducted in heterogenous manners. For instance, some of the studies were conducted with student populations [29,33,41]. Second, the materials and procedures varied significantly between studies: while most asked participants to evaluate storyboards illustrating each mechanism [26,28,37–39], and one study involved participants selecting mechanisms they would prefer to use within a mobile application [42]. Finally, the health-related issues addressed by the studies were heterogeneous, ranging from unhealthy alcohol consumption [37–39] to physical activity [28] and mental health [27,29], further limiting direct comparison.

## Conclusions

Mobile apps represent an intriguing avenue for facilitating the adoption of healthier behaviors among individuals. To optimize the efficacy of these applications in influencing behavior, it is recommended that the content of the app be tailored to the specific profile of the user. This study permitted the delineation of diverse profiles, including those pertaining to personality and gamer profiles, with the Big Five and Hexad scale exhibiting the greatest number of associations. The preferred mechanisms for each of these profiles were then specified. Nevertheless, evidence was found for only 45% of the potential relations. Of note, several relations were identified in the domains of competition, collectibles, progression, customization and cooperation. To experimentally validate the findings of this scoping review, it would be valuable to conduct a study wherein participants’ profiles are measured and they are subsequently asked to select their preferred mechanisms. Such an experiment would serve to corroborate the identified relations and explore any missing connections.

## Data Availability

Data sharing is not applicable to this article as no datasets were generated or analyzed during the study.

## Acknowledgments

LG carried out the scoping review with the help and advice of FE and GF. All authors participated in the writing and reading of the manuscript and approved the final version.

